# Composition of the immunoglobulin G glycome associates with the severity of COVID-19

**DOI:** 10.1101/2020.10.17.20214205

**Authors:** Tea Petrović, Inês Alves, Dario Bugada, Julio Pascual, Frano Vučković, Andrea Skelin, Joana Gaifem, Judit Villar-Garcia, Manuel M. Vicente, Ângela Fernandes, Ana M. Dias, Ivan-Christian Kurolt, Alemka Markotić, Dragan Primorac, Adriana Soares, Luis Malheiro, Irena Trbojević-Akmačić, Miguel Abreu, Rui Sarmento e Castro, Silvia Bettinelli, Annapaola Callegaro, Marco Arosio, Lorena Sangiorgio, Luca F Lorini, Xavier Castells, Juan P. Horcajada, Salomé S. Pinho, Massimo Allegri, Clara Barrios, Gordan Lauc

## Abstract

A large variation in the severity of disease symptoms is one of the key open questions in COVID-19 pandemics. The fact that only a small subset of people infected with SARS-CoV-2 develop severe disease suggests that there have to be some predisposing factors, but biomarkers that reliably predict disease severity have not been found so far. Since overactivation of the immune system is implicated in a severe form of COVID-19 and the IgG glycosylation is known to be involved in the regulation of different immune processes, we evaluated the association of inter-individual variation in IgG N-glycome composition with the severity of COVID-19. The analysis of 166 severe and 167 mild cases from hospitals in Spain, Italy and Portugal revealed statistically significant differences in the composition of the IgG N-glycome. The most notable difference was the decrease in bisecting *N*-acetylglucosamine (GlcNAc) in severe patients from all three cohorts. IgG galactosylation was also lower in severe cases in all cohorts, but the difference in galactosylation was not statistically significant after correction for multiple testing. To our knowledge, this is the first study exploring IgG N-glycome variability in COVID-19 severity.

## Introduction

The knowledge about SARS-CoV-2 virus and COVID-19 increased tremendously in the last few months, but it is still unclear why some infected patients have a very mild disease, or even no symptoms, while others develop the serious disease with considerable mortality. Over a million COVID-19 related deaths have been reported so far, but a significant number of people (even over 80% in some populations) infected with SARS-CoV-2 manage to contain infection only to their upper respiratory tract and despite being positive for the virus do not develop any visible symptoms (Oran & Topol, 2020). Many factors have been raised as clinical predictors of worse disease progression, such as obesity, diabetes, hypertension, kidney injury and in general, the previous cardiovascular disease burden of the patient (Williamson et al., 2020). Undoubtedly, age is the most important factor predicting severity with 100-fold difference in mortality risk in different age groups (Sehra et al., 2020). Several independent studies suggested that environmental factors (temperature and humidity) play an important role for viral transmission, but also for severity of the disease (Kifer et al., 2020; Lauc et al., 2020).

Over-activation of the immune system and the subsequent “cytokine storm” have been implicated in the severe form of COVID-19 (Domingo et al., 2020), but biomarkers that could help identify people at high risk are not available. The first biomarkers offering some insight into inter-individual differences in response to COVID-19 infection were the ABO blood groups, with blood group A individuals having approximately 25% higher risk to get infected with SARS-CoV-2 (Wu et al., 2020). This was subsequently confirmed by a genetic study that identified ABO gene as one of the main genetic risk factors with similar odds ratios (1.45 for blood group A and 0.65 for blood group O) (Ellinghaus et al., 2020). Blood protein profiling on 59 COVID-19 cases and 28 controls using the OLINK inflammation, autoimmune, cardiovascular and neurology panels identified six proteins (IL6, CKAP4, Gal-9, IL-1ra, LILRB4 and PD-L1) that associated with disease severity (Patel et al., 2020). In another small study proteomic and metabolomic profiling of sera from 46 COVID-19 and 53 control individuals (Shen et al., 2020) identified characteristic protein and metabolite changes in the sera of severe COVID-19 patients, which might be used in the selection of potential blood biomarkers for severity evaluation. A recent study suggested that serum CCL17 may be a predictive marker to distinguish between mild/moderate and severe/critical disease in patients with COVID-19 (Sugiyama et al., 2021). Furthermore, it has been shown that patients with severe forms of SARS-CoV-2 pneumonia had neutralizing Immunoglobulin G (IgG) autoantibodies against type I IFNs. Thus, inter-individual variability in IgG could help to explain the immense clinical variability of the infection response (Bastard et al., 2020).

Immunoglobulin G (IgG) glycome composition is an essential component of the immune system that regulates inflammation at multiple levels (Nimmerjahn & Ravetch, 2008; Seeling et al., 2017) and is considered to be one of the important drivers of inflammaging (Franceschi et al., 2018). Many observational and molecular studies of the IgG glycome identified and confirmed its role of both a biomarker and a functional effector of inflammation that contributes to the development of different inflammatory diseases (Lauc et al., 2016). IgG glycome composition has not yet been thoroughly addressed in COVID-19 infection, especially in the context of disease severity. A recent (still unreviewed) small study suggested that the generation of afucosylated antigen-specific IgG may be an important element in the defense against SARS-CoV-2 and other enveloped viruses (Larsen et al., 2020). In another unreviewed study antigen-specific IgG Fc fucosylation in PCR-diagnosed COVID-19 patients was reduced compared to SARS-CoV-2-seropositive children and relative to adults with symptomatic influenza virus infections (Chakraborty et al., 2020). To address this question, we analysed the total IgG N-glycome composition in three independent cohorts of COVID-19 patients.

## Results

IgG glycome composition was analyzed in 167 patients with mild and 166 patients with severe form of COVID-19 from three independent cohorts. The descriptive information about included patients is presented in Table 1. Total IgG N-glycome (combined Fc and Fab glycans) composition was determined by ultra-high-performance liquid chromatography (UHPLC) analysis of glycans labelled with2-aminobenzamide (2-AB) as described in the Materials and methods section.

**Table 1.** Descriptive information about COVID-19 patients included in the study.

|  | Italy (n = 107) |  | Portugal (n = 77) |  | Spain (n = 152) |  |
| --- | --- | --- | --- | --- | --- | --- |
| Disease severity | Mild<br>(n = 40) | Severe<br>(n = 64) | Mild<br>(n = 52) | Severe<br>(n = 25) | Mild<br>(n = 75) | Severe<br>(n = 77) |
| Sex<br>(Male/Female) | 25/15 | 45/19 | 26/26 | 18/7 | 44/31 | 44/33 |
| Age<br>(Median [IQR]) | 70 years<br>(57-76) | 66 years<br>(57-73) | 56 years<br>(44-70) | 64 years<br>(54-81) | 63 years<br>(58-70) | 63 years<br>(55-72) |

Statistical analysis was performed on main summary features of the IgG glycome composition (G0 – glycans without galactose, G1 – glycans with one galactose, G2 – glycans with two galactoses, S – percentage of all glycans with sialic acid, F – percentage of fucosylated glycans, and B – percentage of glycans with bisecting GlcNAc). The analysis of differences between severe and mild cases was performed using a logistic regression model with sex and age included as additional covariates. Significant difference in the IgG glycome composition in severe and mild COVID-19 patients was observed (Table 2, Fig 1). Consistent decrease in the level of bisecting *N*-acetylglucosamine (GlcNAc) in severe cases was observed in all cohorts (meta-analysis effect = - 0.34; adjusted meta-analysis p= 0,009). Galactosylation was also consistently decreased in severe cases in all three cohorts, but the statistical significance of this difference was observed only for monogalactosylation in Barcelona cohort (effect= −0,34; p=0.016). Consistent changes in the levels of sialylated and fucosylated IgG glycan structures between mild and severe COVID-19 cases were not detected.

**Table 2.** IgG glycome composition in severe and mild COVID-19 patients. B – bisecting GlcNAc, G0 – agalactosylation, G1 – monogalactosylation, G2 – digalactosylation, S – sialylation, F - fucosylation.

| **Glycan** | **Italy**<br>**effect** | **Italy**<br>**p val** | **Portugal**<br>**effect** | **Portugal**<br>**p val** | **Spain**<br>**effect** | **Spain**<br>**p val** | **Meta**<br>**effect** | **Meta**<br>**p val** | **Meta**<br>**p adjusted** |
| --- | --- | --- | --- | --- | --- | --- | --- | --- | --- |
| **B**<br>**total** | -0.27 | 0.174 | -0.31 | 0.190 | -0.39 | 0.009 | -0.34 | 0.002 | 0.009 |
| **G0**<br>**total** | 0.14 | 0.437 | 0.12 | 0.547 | 0.21 | 0.101 | 0.17 | 0.065 | 0.112 |
| **G1**<br>**total** | -0.06 | 0.756 | -0.22 | 0.383 | -0.34 | 0.016 | -0.24 | 0.023 | 0.069 |
| **G2**<br>**total** | -0.12 | 0.526 | -0.10 | 0.592 | -0.21 | 0.089 | -0.17 | 0.075 | 0.112 |
| **S**<br>**total** | -0.17 | 0.369 | 0.01 | 0.670 | -0.15 | 0.327 | -0.10 | 0.341 | 0.409 |
| **F**<br>**total** | -0.13 | 0.498 | -0.25 | 0.324 | 0.29 | 0.070 | 0.05 | 0.654 | 0.654 |

**Figure 1:**
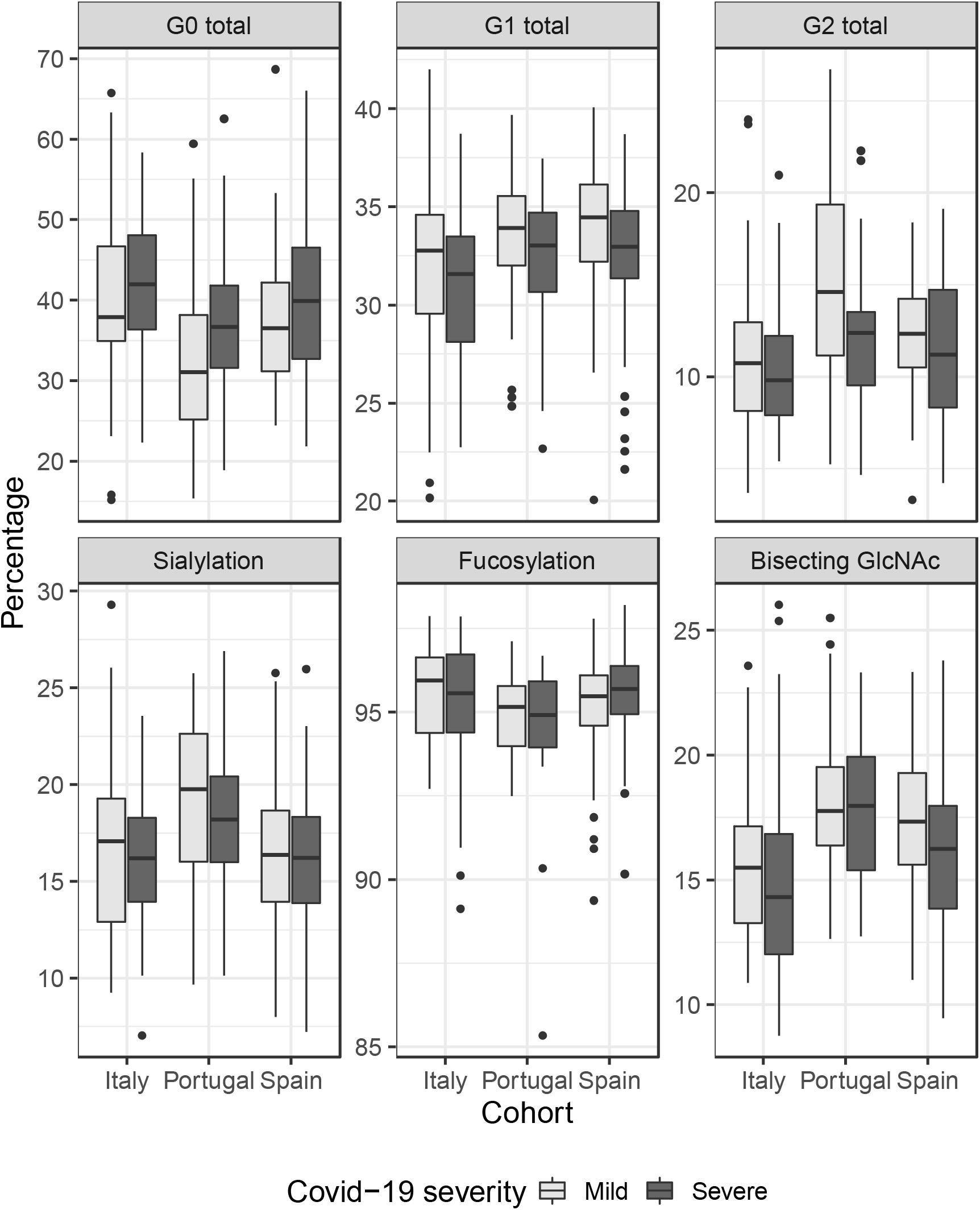
Relative abundance of main IgG glycome features in severe and mild COVID-19 cases from three cohorts. G0 – agalactosylation, G1 – monogalactosylation, G2 – digalactosylation. Boxes represent the 25^th^ and 75^th^ percentiles. Lines inside the box represent the median.

## Discussion

In this study we found consistent differences in IgG glycome composition between patients with mild and severe COVID-19. The level of bisecting GlcNAc was considerably decreased in severe patients (effect = −0.34), replicated in all three cohorts. Despite the small size of each cohort, this change was statistically significant even after adjusting for multiple testing (adjusted meta-analysis p = 0,009). Higher levels of bisecting GlcNAc on IgG are often associated with increased FcγRIII binding and enhanced antibody-dependent cell cytotoxicity (ADCC), explaining a more pro-inflammatory effector functions of IgGs (Irvine & Alter, 2020; Umana et al., 1999). This study had a cross-sectional design that does not allow us to distinguish whether the observed associations reflect a pre-existing risk factor, or rapid changes in IgG glycosylation that occurred during the disease. This question will be addressed in a longitudinal study that we are currently setting up. A recent small study on antigen-specific antibodies found a decrease in bisecting GlcNAc specifically on anti-SARS-CoV2 antibodies (Larsen et al., 2020), which is consistent with changes in the total IgG glycome observed in our study. Interestingly, in the same study anti-SARS-CoV2 antibodies had higher galactosylation and sialylation (relative to the total IgG glycome composition), which is the opposite of what we have observed in this study. This difference suggests that the observed differences in IgG glycosylation between mild and severe cases may not be not a simple reflection of the increased proportion of newly synthetised anti-SARS-CoV2 antibodies in the total IgG pool. However, another study found that IgG1 against the receptor binding domain of the SARS-CoV-2 spike protein from COVID-19 patients had significantly lower core fucosylation, galactosylation and bisection when compared with total IgG1 from healthy adult controls (Chakraborty et al., 2020), suggesting that these changes/differences may be very individual and that additional studies are needed.

“Cytokine storm” and over-activation of the immune system have been suggested key features of severe COVID-19 (Catanzaro et al., 2020), but this significantly varies from patient to patient (Calfee et al., 2014) and increased levels of pro-inflammatory cytokines are not present in all severe patients (Kox et al., 2020). Inter-individual differences in IgG glycosylation are large and reflect both genetic and environmental contributing factors (Klarić et al., 2020; Pučić et al., 2011). IgG glycans are known effectors of the immune system and composition of the IgG glycome associates with different diseases (Gudelj et al., 2018).

It has been shown that age and excess adiposity is a risk factor for severe disease and mortality in people with SARS-CoV-2 infection (Seidu et al., 2020). Furthermore, age (Krištić et al., 2014) and adiposity (Russell et al., 2019) are the main environmental factors that drive the decrease in IgG glycosylation, so it is intriguing to hypothesize that changes in the IgG glycome that lead to the loss of its immunosuppressive potential may be one of the molecular mechanisms behind these two environmental risk factors for severe COVID-19. IgG glycome composition strongly associates with age, so it is very hard to exclude confounding factors with some other age-related changes, but the fact that people with severe COVID-19 had “older” IgG glycome composition suggests the need for further research in this direction.

## Materials and Methods

### Studied cohorts

Biological samples were obtained from hospitals in Spain, Italy, and Portugal. The study protocol conformed to the ethical guidelines of the 1975 Declaration of Helsinki and the study was approved by ethical committees of ASST Papa Giovanni XXIII° Hospital in Bergamo, Hospital del Mar in Barcelona and by the institutional ethics committee of Centro Hospitalar Universitário do Porto (CHUP), Centro Hospitalar de Vila Nova de Gaia/Espinho (CHVNG) and Hospital Beatriz Angelo (HBA), Loures, Lisbon. All participants gave informed consent.

#### Spain

Parc de Salut MAR Biobank (MARBiobanc) Barcelona. Patients with PCR confirmed SARS-CoV-2 infection admitted at the Hospital del Mar in Barcelona during the months of March-May were included in this study We classified the patients in severe or mild COVID, considered severe if they needed invasive or non-invasive mechanical ventilation or intensive care unit, or deceased during the hospitalization.

#### Italy

ASST Papa Giovanni XXIII° Hospital, Bergamo. Hospitalized patients for acute respiratory failure due to SARS-CoV-2 infection were included in this study. They were divided in two categories: a) *severe* disease: all patients requiring an ICU admission for respiratory failure, and treated with either mechanical ventilation or CPAP (Continuous Positive Airways Pressure) and b) *mild* disease: all patients who required oxygen support by mask or no support at all.

#### Portugal

Patients from Infectious Disease Department of CHUPCHVNG and HBA were included. Blood was collected for plasma at the time of diagnosis. SARS-CoV-2 positive patients with different levels of severity (mild, moderate and severe) were include. The WHO criteria were used to stratify symptomatic SARS-CoV-2 patients by the disease severity into mild: individuals with no evidences of pneumonia; moderate: individuals with evidence of pneumonia, however without need of invasive mechanical ventilation and without need of admission to hospital intensive care unit; severe: individuals with need of invasive mechanical ventilation and with need of admission to hospital intensive care unit. For this study moderate and severe patients were merged into a single group.

### Isolation of IgG from human plasma

IgG was isolated from plasma using a 96-well protein G monolithic plate (BIA Separations, Slovenia). Before use the monolithic plate was preconditioned with ultrapure water, 10x PBS, pH 6.6, 0.1 M formic acid, pH 2.5 (Honeywell Fluka, UK) and 1 × PBS, pH 7.4. 100 µl of plasma was diluted 8 × 1 × PBS, pH 7.4, applied to the Protein G plate and washed with 1x PBS, pH 7.4 to remove unbound proteins. IgG was eluted from monolithic plate with 0.1 M formic acid, pH 2.5. Eluates were collected in a 96-deep-well plate and neutralized with 1M ammonium bicarbonate (Acros Organic, USA). After each sample application, the monoliths were regenerated with the following buffers: 0.1 M formic acid pH 2.5, followed by 10 × PBS acid and afterward 1 × PBS to re-equilibrate the monoliths. Each step was done under vacuum (cca. 10 inHg pressure reduction during sample application and IgG elution, 17 inHg during elution and washing steps) using a manual set-up consisting of a multichannel pipette, a vacuum manifold (Pall Corporation, USA) and a vacuum pump (Pall Corporation, USA). The monolithic plate was stored in 20 % (v/v) EtOH in 20 mM TRIS + 0.1 M NaCl, pH 7.4 at 4 °C.

### Glycan release, labelling and clean up

After IgG isolation, IgG eluates were heated at 65 °C for 30 minutes before drying to reduce risk from any potential residual virus in the IgG eluate. Samples were dried in a vacuum concentrator and denatured in 30 µL 1.33% SDS (w/v) (Sigma-Aldrich, USA) by incubation at 65 °C for 10 minutes. After cooling to room temperature for 30 minutes, 10 µL of 4% Igepal-CA630 (Sigma-Aldrich, USA) were added to the samples and incubated on a shaker for 15 minutes. For glycan release, 1.25 mU of PNGase F (Promega, USA) in 10 µL 5× PBS were added and incubated for 18 hours at 37 °C.

The released *N*-glycans were labelled with 2-AB (Sigma-Aldrich, USA). The labelling mixture was freshly prepared by dissolving 2-AB (Sigma-Aldrich, USA) and 2-picoline borane (Sigma-Aldrich, USA) in dimethylsulfoxide (DMSO) (Sigma-Aldrich, USA) and glacial acetic acid (Honeywell Fluka, UK) mixture (70 : 30 v/v) to the final concentration of 19,2 mg/mL for 2AB and 44,8 mg/mL for 2-picoline borane. To each *N*-glycan sample in the 96-well plate, 25 µl of labelling mixture was added, and the plate was sealed using an adhesive seal. Mixing was achieved by shaking for 10 min, followed by 2 h incubation at 65°C. The reaction mixture was cooled down to room temperature for 30 minutes. Samples (75 µL) were diluted with 700 µL of Acetonitrile (Carlo Erba, Spain) prior to clean up procedure by hydrophilic interaction liquid chromatography solid-phase extraction (HILIC-SPE).

As a stationary phase, 0.2 µm Supor AcroPrep filter plate (Pall Corporation, USA) was used and wells were prewashed with 200 µL 70% ethanol (v/v), 200 µL ultrapure water and 200 µL 96% cold ACN (v/v) (Carlo Erba, Spain). The samples were loaded into the wells, and after a short incubation washed with 5 × 200 µl 96% ACN (Carlo Erba, Spain). Glycans were eluted with 2 × 90 µl of ultrapure water after 15 min shaking at room temperature, and combined eluates were stored at –20°C until use.

### Hydrophilic Interaction Liquid Chromatography (HILIC)-UHPLC

Fluorescently labelled N-glycans were separated by hydrophilic interaction chromatography on a Waters Acquity UHPLC instrument (Milford, MA, USA). The instrument consists of a sample manager, a quaternary solvent manager and a FLR fluorescence detector. Released and labelled N-glycans were chromatographically profiled by HILIC using an amide-bonded sub 2 micron stationary phase column built from a novel type of column hardware (ACQUITY PREMIER Glycan BEH Amide 130 Å, 1.7 µm 2.1 x 100 mm Column, Waters Corporation, Milford, MA). Unlike traditional, metallic hardware, this column was constructed from components manufactured to have a barrier layer of hybrid organic inorganic silica to improve its inertness and to mitigate problematic analyte to metal adsorption. Having a chemical composition that is highly similar to ethylene-bridged siloxane (BEH) particles (D. Wyndham et al., 2003), this barrier layer is considerably more inert than fused silica surfaces and significantly less hydrophobic than polyether ether ketone (PEEK). That it is comprised of highly crosslinked ethylene-bridged siloxane groups also means that it is resilient to chemical and pH stress and amenable to usage between pH 1 and pH 12 mobile phase conditions.

Separation method used a linear gradient of 75–62% acetonitrile (v/v) at a flow rate of 0.4 ml/min in a 29 minutes analytical run. Separation temperature was 60 °C, and samples were maintained at 10 °C before injection. Hydrolysed and 2-AB labelled glucose oligomers were used as external standard, from which the retention times for the individual glycans were converted to glucose units. The chromatograms were separated into 24 chromatographic peaks and the amount of glycans in each peak was expressed as percentage of total integrated chromatogram area (% Area).

### Statistical analysis

In order to remove experimental variation from measurements, normalization and batch correction were performed on UHPLC glycan data. To make measurements across samples comparable, normalization by total area was performed where the peak area of each of 24 glycan structures was divided by the total area of the corresponding chromatogram. Prior to batch correction, normalized glycan measurements were log transformed due to right-skewness of their distributions and the multiplicative nature of batch effects. Batch correction was performed on log-transformed measurements using ComBat method (R package sva), where the technical source of variation (which sample was analyzed on which plate) was modeled as a batch covariate. To get measurements corrected for experimental noise, estimated batch effects were subtracted from log-transformed measurements. In addition to 24 directly measured glycan structures, six derived traits were calculated from the directly measured glycans. These derived traits average particular glycosylation features across different individual glycan structures and consequently they are more closely related to individual enzymatic activities and underlying genetic polymorphisms.

Association analyses between disease severity status and glycan traits were performed using a regression model with age and gender included as additional covariates. Analyses were firstly performed for each cohort separately and then combined using inverse-variance weighted meta-analysis approach (R package metafor). Prior to analyses, glycan variables were all transformed to standard Normal distribution (mean=0, sd=1) by inverse transformation of ranks to Normality (R package “GenABEL”, function rntransform). Using rank transformed variables in analyses makes estimated effects of different glycans in different cohorts comparable as transformed glycan variables have the same standardized variance. False discovery rate was controlled using Benjamini-Hochberg procedure. Data was analysed and visualized using R programming language (version 3.0.1).

## Data Availability

Data is available from corresponding authors

## Funding

This work was supported in part by the European Structural and Investment Funds grant for the Croatian National Centre of Competence in Molecular Diagnostics (#KK.01.2.2.03.0006), National Centre of Research Excellence in Personalized Healthcare grant (#KK.01.1.1.01.0010) and by Croatian Science Foundation project #IP CORONA-2020-04. MARBiobanc’s work was supported by grants from Instituto de Salud Carlos III/FEDER (PT17/0015/0011) and the “Xarxa de Bancs de tumors” sponsored by Pla Director d’Oncologia de Catalunya (XBTC). Clara Barrios is funded by grants FIS-FEDER-ISCIII PI16/00620 and PERIS STL008.

The Institute of Molecular Pathology and Immunology of the University of Porto integrates the Institute for Research and Innovation in Health (i3S) research unit, which is partially supported by the Portuguese Foundation for Science and Technology (FCT). This article was funded by the FCT, in collaboration with the Portuguese Agency for Clinical Research and Biomedical Innovation (AICIB) under the special funding, “RESEARCH 4 COVID-19” Project #006 granted to the PI Salomé S. Pinho. This article is also a result of the project NORTE-01-0145-FEDER-000029, supported by the Norte Portugal Regional Programme (NORTE 2020) under the PORTUGAL 2020 Partnership Agreement through the European Regional Development Fund. This work was also funded by Fundo Europeu de Desenvolvimento Regional (FEDER) funds through the COMPETE2020—Operacional Programme for Competitiveness and Internationalization (POCI), Portugal 2020, and by Portuguese funds through the FCT in the framework of the project (POCI-01-0145-FEDER-028772).

IA [SFRH/BD/128874/2017] and MV [PD/BD/135452/2017] received funding from the FCT.

## Acknowledgements

We would like to thank all anesthesiologists and nurses in the Emergency and Intensive Care Unit, as well as all the people who worked at ASST Papa Giovanni XXIII, during the COVID-19 pandemic in Bergamo for their invaluable and brave efforts towards patient’s care. We would like to thank the ROCCO Bergamo project for its scientific support. Furthermore, we want to particularly acknowledge the patients of the Parc de Salut Mar and the MARBiobanc, integrated in the Spanish National Biobanks Network, for their collaboration. We thank Waters Corporation, USA, for supporting this work by providing ACQUITY PREMIER Glycan BEH Amide columns.

We would like to thank to the clinicians Dr. Tiago Teixeira, Dr. Sónia Rocha, Dr. Sofia Nunes, and Dr. Cristóvão Figueiredo from CHVNG and to Dr. Ana Rita Silva, Dr. Salomão Fernandes, Dr. Bernardo Silva and Dr. Rita Sérvio from HBA that were involved in COVID-19 patients’care and provided support in patients recruitment for this study. We would like also to thank to all the patients that accepted to participate in this study, as well as to the nurses and technicians that collaborated in the collection of the samples including Nurse Teresa Cruz from CHUP, Dr. Nair Seixas from CHVNG and the director of Clinical Pathology Lab from CHUP, Dr. José Carlos Oliveira for all the support.

## Abbreviations

2-AB: 2-aminobenzamide
B: percentage of glycans with bisecting
GlcNAcBEH: ethylene-bridged siloxane
CFR: case fatality rates
DMSO: dimethylsulfoxide
F: percentage of fucosylated glycans
G0: percentage of glycans without galactose
G1: percentage of glycans with one galactose
G2: percentage of glycans with two galactoses
GlcNAc: N-acetlyglucosamine
HILIC: hydrophilic interaction liquid chromatography
IgG: immunoglobulin G
PEEK: polyether ether ketone
S: percentage of all glycans with sialic acid
SPE: solid-phase extraction
UHPLC: ultra-high-performance liquid chromatography

## Notes

### Competing Interest Statement

The authors have declared no competing interest.

### Clinical Trial

This was a retrospective study on leftover diagnostic samples

### Author Declarations

Biological samples were obtained from hospitals in Spain, Italy, and Portugal. The study protocol conformed to the ethical guidelines of the 1975 Declaration of Helsinki and the study was approved by ethical committees of ASST Papa Giovanni XXIII Hospital in Bergamo, Hospital del Mar in Barcelona and by the institutional ethics committee of Centro Hospitalar Universitario do Porto (CHUP), Centro Hospitalar de Vila Nova de Gaia/Espinho (CHVNG) and Hospital Beatriz Angelo (HBA), Loures, Lisbon. All participants gave informed consent.

